# Trends and distribution of birth asphyxia, Uganda, 2017-2020: A retrospective analysis of public health surveillance data

**DOI:** 10.1101/2023.01.09.23284352

**Authors:** Allan Komakech, Freda L. Aceng, Stella M. Migamba, Petranilla Nakamya, Robert Mutumba, Lilian Bulage, Benon Kwesiga, Alex R. Ario

## Abstract

**Background:** During 2018-2020, almost half of all neonatal deaths reviewed in Uganda were due to birth asphyxia. In 2015, Uganda adopted the Every Newborn Action Plan interventions to renew focus on surveillance for birth asphyxia and other childhood-related illnesses. In 2016, Ministry of Health implemented an evidence-based educational program for birth attendants about neonatal resuscitation techniques to improve management of birth asphyxia. Birth asphyxia is reported on a monthly basis in Uganda as part of routine reporting. We described the trends and distribution of birth asphyxia in Uganda during 2017–2020 following these renewed efforts.

**Methods:** We analysed birth asphyxia surveillance data from the District Health Information System 2 during January 2017–December 2020. We calculated incidence of birth asphyxia per 1,000 deliveries at district, regional, and national levels. We used line graphs to demonstrate the trend of birth asphyxia incidence with the corresponding reporting rates at national and regional levels and used logistic regression to evaluate significance of the trends. Using choropleth maps, we described the distribution of birth asphyxia incidence at district level.

**Results:** The average national annual incidence of birth asphyxia increased by 4.5% from 2017 to 2020 (OR=1.05; 95%CI=1.04-1.05, p=0.001), with national quarterly reporting rates of 70-80% over the same period. Incidence in the Northern and Eastern Regions increased 6% (OR=1.06; 95%CI=1.05-1.07, p=0.001) and 5% (OR=1.05; 95%CI=1.03-1.05, p=0.001), respectively, over the study period. Bundibugyo, Iganga, and Mubende Districts had rates of >60/1,000 during each of the four years of the study period. The least affected district was Kazo District, with an overall incidence of 3/1,000 over the study period.

**Conclusion:** The incidence of birth asphyxia increased nationally from 2017-2020. Continuous capacity-building in birth asphyxia management, with emphasis on the most affected districts, could reduce the burden of this public health problem in Uganda.

## Introduction

The World Health Organization (WHO) defines birth asphyxia as the failure to initiate and sustain breathing at birth (1). Birth asphyxia is a leading cause of brain damage among newborn children, with up to 80% of survivors suffering from disabilities, developmental delays, palsy, intellectual disabilities, and behavioural problems (2–4) Worldwide, birth asphyxia is responsible for an estimated 900,000 neonatal deaths per year (5). Birth asphyxia incidence in developing countries has previously been reported to be 10 times that in developed countries (6). Studies done in Bangladesh (7) and in Nigeria (8) observed that birth asphyxia was responsible for 39% and 23.3%, respectively, of all neonatal deaths.

Risk factors for birth asphyxia are grouped according to whether they occur before birth (antepartum risk factors), during birth (intrapartum risk factors), or after birth (postpartum risk factors). Antepartum risk factors include severe maternal hypotension or hypertensive diseases during pregnancy, history of stillbirth, young maternal age, and advanced maternal age. Intrapartum risk factors include malpresentation of the fetus, a prolonged second stage of labour, and home delivery. Postpartum risk factors include low birth weight, high birth weight, preterm delivery, and poor resuscitation efforts (9–12). The majority of these are preventable, as evidenced by the regional variations across the world

During 2018-2020, almost half of all neonatal deaths reviewed in Uganda were due to birth asphyxia (13). Studies done in Uganda implicated antepartum and intrapartum risk factors, including complexities of referral systems, non-attendance of antenatal care by mothers, knowledge gaps among health workers, lack of equipment and high health worker-to-patient ratio as the major culprits (14–16). In 2015, Uganda adopted the Every Newborn Action Plan (17), which included a renewed focus on surveillance of birth asphyxia cases and development of a national strategic plan on managing birth asphyxia and other childhood illnesses. Furthermore, in 2016 the Ministry of Health rolled out the Helping Babies Breathe (HBB) initiative, meant to improve prevention and management of birth asphyxia by health workers. However, the impact of these interventions on birth asphyxia incidence was unknown. We described the trends and distribution of birth asphyxia in Uganda during 2017-2020, the era following these renewed efforts.

## Materials and Methods

### Study setting

Uganda is located in East Africa with an estimated population of 41.6 million people (18). There are four regions in Uganda, each of which is partitioned into 135 districts and 11 cities (18). The health system comprises public and private health sectors and healthcare is provided through a decentralized system in which services are delivered across seven tiers. These include national referral hospitals, regional referral hospitals, district hospitals, Health Centres IV (HCIV), Health Centres III (HCIII), Health Centres II (HCII), and community health workers locally referred to as the Village Health Teams (VHTs) (19). In Uganda, birth asphyxia is a condition that can be managed at HCII facilities and above.

### Study design and data source

We conducted a nationwide surveillance data analysis of birth asphyxia cases from 2017 to 2020 using data abstracted from the Uganda District Health Information System 2 (DHIS2), a web-based reporting tool introduced to Uganda in 2012. Data on birth asphyxia data are routinely generated at health facilities using the integrated maternity register.

The data from these registers are aggregated into a health facility monthly report (paper form) which is submitted to health sub-district and then to the district health offices. At the district health office, data from the paper-based reports are entered into DHIS2. Data in DHIS2 are then grouped at national, regional, district, sub-county, and facility levels. At the national level, the Reproductive and Infant Health Department of the Ministry of Health and other stakeholders use data from DHIS2 to make decisions and plan interventions on reproductive and infant health.

### Study variables, data abstraction and analysis

We calculated national quarterly reporting rates of birth asphyxia by dividing the available monthly reports per quarter for all districts by the expected monthly reports for all districts per quarter. We abstracted data for birth asphyxia cases and total deliveries during 2017–2020 from the DHIS2. We disaggregated the data into national, regional, and district levels. We calculated the annual incidence rates per 1,000 for birth asphyxia cases at national, regional, and district levels by dividing the total birth asphyxia cases during the year by the total deliveries during that year and multiplying by 1,000. We obtained the mean annual incidence rates for the national and regional levels by adding annual incidence rates for the four years of study and dividing by four.

We plotted the quarterly mean incidence of birth asphyxia against the study period in years to present the trend in incidence at national and regional levels from 2017–2020 and used logistic regression to establish the significance of the trends. We also used choropleth maps generated using Quantum Geographic Information System (QGIS) to present the district distribution of the birth asphyxia incidence across the country.

## Results

### Trend of annual incidence rate of birth asphyxia, national level, Uganda, 2017–2020

In Uganda, there were a total of 4,625,336 deliveries and 134,801 birth asphyxia cases during 2017-2020. The average national incidence of birth asphyxia over the four years in Uganda was 29 per 1,000 deliveries. The highest annual incidence (32 birth asphyxia cases per 1,000 deliveries) over the four years was recorded in 2020.

The incidence of birth asphyxia increased from 2017–2020 (OR 1.045; 95% CI 1.04, 1.05) **(Figure 1)**. Reporting rates remained fairly stable (between 70% and 80%) from January 2017 to December 2020.

**Figure 1:**
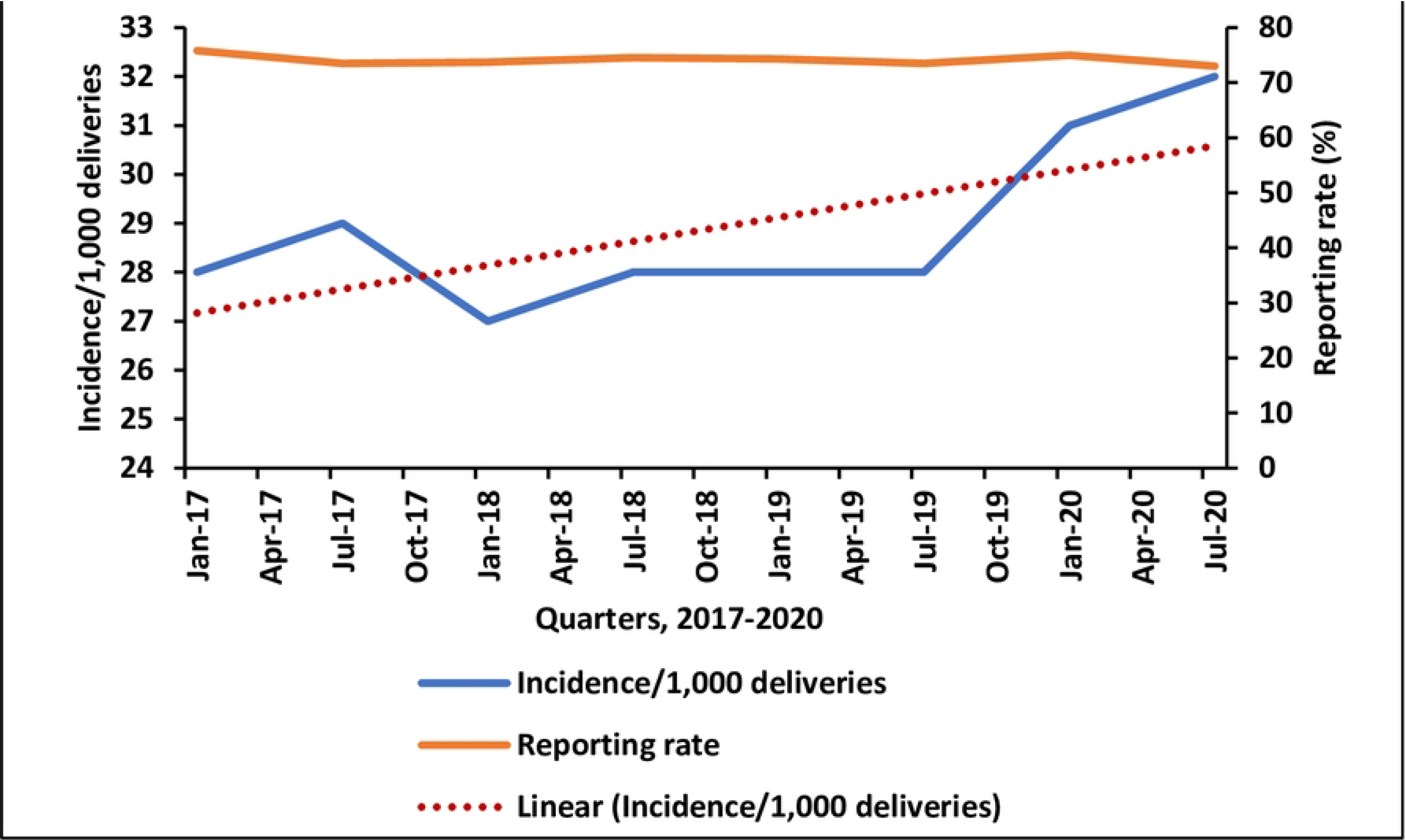
Quarterly trends of birth asphyxia incidence/1,000 total deliveries, Uganda, 2017– 2020.

### Trend of annual incidence rate of birth asphyxia cases, regional level, Uganda, 2017-2020

The increase in the incidence rates of birth asphyxia/1,000 total deliveries were limited to the Northern (7%) and Eastern (5%) regions of Uganda, while rates in other regions remained unchanged **(Table 1)**. Central Region registered the highest mean annual incidence rate of 30 birth asphyxia cases/1,000 total deliveries and Northern Region registered the lowest mean incidence rate of 28 per 1,000 deliveries over the four years.

**Table 1:** Significance of trends of birth asphyxia incidence at regional level, Uganda, 2017–2020.

| Region 2017/2020 | Odds Ratio | 95% CI | P-Value |
| --- | --- | --- | --- |
| Northern | 1.07 | 1.05-1.08 | <0.001* |
| Eastern | 1.05 | 1.04-1.06 | <0.001* |
| Central | 1.00 | 0.99-1.01 | 0.54 |
| Western | 0.99 | 0.99-1.01 | 0.75 |

\*indicates p value<0.05

### Spatial distribution of birth asphyxia incidence rates, district level, Uganda, 2017-2020

There was minimal spatial clustering of high-incidence districts for birth asphyxia during 2017-2020. However, specific districts had persistently high birth asphyxia incidence rates over the study period; these included Bundibugyo, Mubende, and Iganga **(Figure 2**).

**Figure 2:**
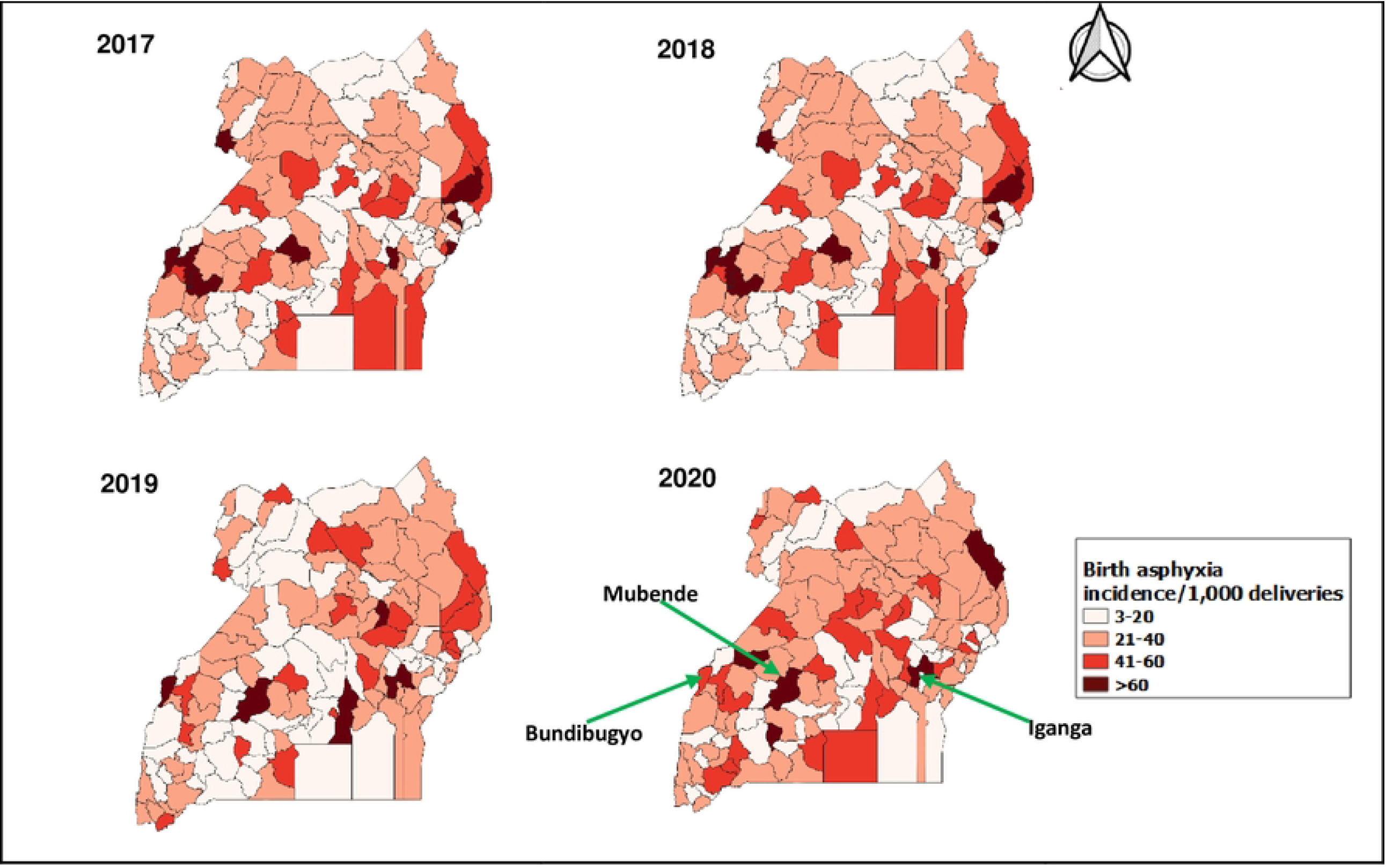
Spatial distribution of birth asphyxia cases/1,000 total deliveries, Uganda, 2017–2020.

## Discussion

Birth asphyxia incidence rates increased significantly in Uganda during 2017-2020, specifically due to increases in the Northern and Eastern Regions. Spatial trends showed minimal clustering of birth asphyxia incidence in the country. However, specific districts faced persistently high birth asphyxia incidence rates over the four years.

While the reasons for the increase in birth asphyxia incidence over the study period is not clearly understood, the burden of birth asphyxia is particularly high in East and Central Africa compared to other regions of Sub-Saharan Africa (20,21) This is due to poor obstetrics coverage, inequity and inequality resulting from gaps in local health financing models, inaccessible health facilities, socio-cultural norms, low literacy levels, shortage in health workers and supplies, low level of health service utilization and inadequate resource allocation to healthcare (21–23). Funding health services can lead to provision of services vital to reduce and prevent birth asphyxia. The highest incidence of birth asphyxia during the study period occurred in 2020 at 32 birth asphyxia cases per 1,000 deliveries compared to 28/1000 total deliveries in the three previous years. The is likely due to the delayed access of pregnant women to health facilities following the imposition of the COVID-19 lockdown travel restrictions (24). A study in a rural hospital in Central Uganda showed an increase of 7% in birth asphyxia incidence during the early phase of the COVID-19 lockdown versus before the lockdown (25). The COVID-19 pandemic caused massive disruptions throughout many Africa countries affecting access to health facilities as evidenced in studies in Malawi, Ethiopia and Ghana (26–28). Furthermore, crisis situations do not necessarily lead to reduction in reproduction and yet access to health services is greatly affected during such periods. Special considerations should therefore be ensured to facilitate the access of pregnant women to health facilities during lockdown situations to improve access to health care.

While several districts had intermittently high rates of birth asphyxia, Bundibugyo, Iganga, and Mubende districts had persistently high incidences of >60 cases per 1,000 deliveries. This is approximately twice the national average rate. Kazo District was the least affected district with 3 cases of birth asphyxia/1,000 deliveries. Although no particular reason can be identified to explain the non-clustered patterns of birth asphyxia in Uganda, all regions of Uganda have their fair share of poverty (29), issues with access to- and availability of health facilities (30), and differing cultural and social norms, known factors that can lead to birth asphyxia. Studies done to establish socio-demographic and health facility factors associated with birth asphyxia, particularly in the highly affected regions would be beneficial.

## Limitations

Secondary data in DHIS2 is limited in terms of variables to provide a sufficient assessment of birth asphyxia incidence in Uganda. Studies using primary data to determine associated factors may be more beneficial in understanding increasing birth asphyxia trends in Uganda. This will help to improve already-existing evidence-based interventions. Secondly, given the low average reporting rates over the study period (<80%), a true representation of birth asphyxia incidence might be limited. It should also be noted that since some deliveries occur outside the hospital, it is possible that the incidence rates we are reporting here are an underestimate.

## Conclusion

Birth asphyxia incidence increased over the four years of our study period despite a decrease in reporting rates. The highest incidence over the four years was recorded in 2020. Bundibugyo, Iganga, and Mubende districts had a persistently high birth asphyxia incidence (>60/1,000 deliveries). Kazo district was the least affected district (3/1000 deliveries).

We recommend the Ministry of Health to emphasize consistent reporting of birth asphyxia to ensure useful surveillance data. We also recommend continuous capacity building in managing birth asphyxia, with emphasis on the most affected districts.

## Data Availability

The datasets used for this descriptive analysis are a property of the Uganda MoH and are not publicly available. The data is stored in a password protected database and used for making decisions at a national level. However, with a reasonable request and permission from Uganda MoH, data can be accessed through the corresponding authors.

## List of Abbreviations

CI: Confidence Interval
DHIS-2: District Health Information System-version 2
HBB: Helping Babies Breath
MoH: Ministry of Health
OR: Odds Ratio
QGIS: Quantum Geographic Information System
WHO: World Health Organization

## Acknowledgements

We would like to thank the Ministry of Health for providing access to DHIS2 data that was used for this analysis. We appreciate the technical support provided by the Reproductive Health Department of the Ministry of Health. Finally, we thank the US-CDC for supporting the activities of the Uganda Public Health Fellowship Program (UPHFP).

## References

1. World Health Organization (WHO). Newborn Health [Internet]. [cited 2022 Nov 25]. Available from: https://www.who.int/teams/maternal-newborn-child-adolescent-health-and-ageing/newborn-health/perinatal-asphyxia

2. The long-term health, social, and financial burden of hypoxic–ischaemic encephalopathy - Eunson - 2015 - Developmental Medicine & Child Neurology - Wiley Online Library [Internet]. [cited 2022 Dec 9]. Available from: https://onlinelibrary.wiley.com/doi/full/10.1111/dmcn.12727

3. Ahearne CE. Short and long term prognosis in perinatal asphyxia: An update. World J Clin Pediatr [Internet]. 2016 [cited 2022 Dec 9];5(1):67. Available from: https://www.wjgnet.com/2219-2808/full/v5/i1/67.htm

4. Halloran DR, McClure E, Chakraborty H, Chomba E, Wright LL, Carlo WA. Birth asphyxia survivors in a developing country. J Perinatol 2009 293 [Internet]. 2008 Nov 27 [cited 2022 Dec 9];29(3):243–9. Available from: https://www.nature.com/articles/jp2008192

5. World Health Organization (WHO). Newborn Health [Internet]. [cited 2022 Dec 9]. Available from: https://www.who.int/teams/maternal-newborn-child-adolescent-health-and-ageing/newborn-health/perinatal-asphyxia

6. Lawn JE, Lee ACC, Kinney M, Sibley L, Carlo WA, Paul VK, et al. Two million intrapartum-related stillbirths and neonatal deaths: Where, why, and what can be done? Int J Gynecol Obstet. 2009 Oct 1;107(SUPPL.):S5–19.

7. Sampa RP, Hossain QZ, Sultana S. Observation of Birth Asphyxia and Its Impact on Neonatal Mortality in Khulna Urban Slum Bangladesh. Int J Adv Nutr Heal Sci. 2013 Dec 13;1(1):1–8.

8. Adebami OJ. Maternal and fetal determinants of mortality in babies with birth asphyxia at Osogbo, Southwestern Nigeria. Glo Adv Res J Med Med Sci. 2015;4(6):270–6.

9. Aslam HM, Saleem S, Afzal R, Iqbal U, Saleem SM, Shaikh MWA, et al. “Risk factors of birth asphyxia.” Ital J Pediatr 2014 401 [Internet]. 2014 Dec 20 [cited 2021 Jul 22];40(1):1–9. Available from: https://ijponline.biomedcentral.com/articles/10.1186/s13052-014-0094-2

10. Igboanugo S, Chen A, Mielke JG. Maternal risk factors for birth asphyxia in low-resource communities. A systematic review of the literature. https://doi.org/101080/0144361520191679737 [Internet]. 2019 Nov 16 [cited 2021 Jul 22];40(8):1039–55. Available from: https://www.tandfonline.com/doi/abs/10.1080/01443615.2019.1679737

11. Bayih WA, Yitbarek GY, Aynalem YA, Abate BB, Tesfaw A, Ayalew MY, et al. Prevalence and associated factors of birth asphyxia among live births at Debre Tabor General Hospital, North Central Ethiopia. BMC Pregnancy Childbirth 2020 201 [Internet]. 2020 Oct 28 [cited 2021 Jul 22];20(1):1–12. Available from: https://bmcpregnancychildbirth.biomedcentral.com/articles/10.1186/s12884-020-03348-2

12. Panna S. Risk Factors for Birth Asphyxia in Newborns Delivered at Nongkhai Hospital. Srinagarind Med Journal-ศรีนครินทร์เวชสาร [Internet]. 2020 Jun 20 [cited 2021 Jul 22];35(3):278–86. Available from: https://thaidj.org/index.php/SMNJ/article/view/9123

13. Uganda M. FY 2019_2020 Final MPDSR Report for Printing. 2020.

14. Ayebare E, Ndeezi G, Hjelmstedt A, Nankunda J, Tumwine JK, Hanson C, et al. Health care workers’ experiences of managing foetal distress and birth asphyxia at health facilities in Northern Uganda. Reprod Health [Internet]. 2021 Dec 1 [cited 2022 Mar 23];18(1):1–11. Available from: https://reproductive-health-journal.biomedcentral.com/articles/10.1186/s12978-021-01083-1

15. Arach AAO, Tumwine JK, Nakasujja N, Ndeezi G, Kiguli J, Mukunya D, et al. Perinatal death in Northern Uganda: incidence and risk factors in a community-based prospective cohort study. https://doi.org/101080/1654971620201859823 [Internet]. 2021 [cited 2021 Jul 20];14(1). Available from: https://www.tandfonline.com/doi/abs/10.1080/16549716.2020.1859823

16. Ayebare E, Jonas W, Ndeezi G, Nankunda J, Hanson C, Tumwine JK, et al. Fetal heart rate monitoring practices at a public hospital in Northern Uganda – what health workers document, do and say. https://doi.org/101080/1654971620201711618 [Internet]. 2020 Dec 31 [cited 2021 Jul 20];13(1). Available from: https://www.tandfonline.com/doi/abs/10.1080/16549716.2020.1711618

17. World Health Organisation. Every newborn action plan : Country progress tracking report. Who. 2014;58.

18. Uganda Bureau of Statistics (UBOS). uganda profile - Uganda Bureau of Statistics [Internet]. 2022. [cited 2022 Dec 13]. Available from: https://www.ubos.org/uganda-profile/

19. World Health Organization (WHO). HEALTH SITUATION-Uganda. 2018 [cited 2022 Dec 14]; Available from: http://apps.who.int/gho/data/node.cco

20. Kinoti SN. Asphyxia of the newborn in east, central and southern Africa. East Afr Med J [Internet]. 1993 Jul 1 [cited 2022 Dec 14];70(7):422–33. Available from: https://europepmc.org/article/med/8293701

21. Abekah-Nkrumah G. Trends in utilisation and inequality in the use of reproductive health services in Sub-Saharan Africa. BMC Public Health [Internet]. 2019 Nov 21 [cited 2022 Dec 14];19(1):1–15. Available from: https://link.springer.com/articles/10.1186/s12889-019-7865-z

22. Waters D, Adeloye D, Woolham D, Wastnedge E, Patel S, Rudan I. Global birth prevalence and mortality from inborn errors of metabolism: a systematic analysis of the evidence. J Glob Health [Internet]. 2018 [cited 2022 Dec 14];8(2). Available from: /pmc/articles/PMC6237105/

23. Pell C, Meñaca A, Were F, Afrah NA, Chatio S, Manda-Taylor L, et al. Factors Affecting Antenatal Care Attendance: Results from Qualitative Studies in Ghana, Kenya and Malawi. PLoS One [Internet]. 2013 Jan 15 [cited 2022 Dec 14];8(1):e53747. Available from: https://journals.plos.org/plosone/article?id=10.1371/journal.pone.0053747

24. Foundation ER. Sexual and Reproductive Health in Uganda under the Coronavirus Pandemic. 2020.

25. Hedstrom A, Mubiri P, Nyonyintono J, Nakakande J, Magnusson B, Vaughan M, et al. Outcomes in a Rural Ugandan Neonatal Unit Before and During the Early COVID-19 Pandemic: A Retrospective Cohort Study. SSRN Electron J [Internet]. 2021 [cited 2021 Jul 20]; Available from: https://papers.ssrn.com/abstract=3872622

26. Kazanga Chiumia I, Azariah Mosiwa B, Nkhonjera J, Kazanga B, Shingirai Mukondiwa A, Twalibu A, et al. Emerging public health challenges during the COVID-19 pandemic in Malawi: A review. Public Heal Challenges [Internet]. 2022 Dec 1 [cited 2022 Dec 14];1(4):e40. Available from: https://onlinelibrary.wiley.com/doi/full/10.1002/puh2.40

27. Zewdie A, Mose A, Yimer A, Melis T, Muhamed AN, Jemal AK. Essential maternal health service disruptions in Ethiopia during COVID 19 pandemic: a systematic review. BMC Women’s Heal 2022 221 [Internet]. 2022 Dec 6 [cited 2022 Dec 14];22(1):1–8. Available from: https://bmcwomenshealth.biomedcentral.com/articles/10.1186/s12905-022-02091-4

28. Asuming PO, Gaisie DA, Agula C, Bawah AA. Impact of Covid-19 on Maternal Health Seeking in Ghana. J Int Dev [Internet]. 2022 May 1 [cited 2022 Dec 14];34(4):919. Available from: /pmc/articles/PMC9015309/

29. UNICEF. UGANDA’S MULTIDIMENSIONAL POVERTY PROFILE. [cited 2022 Dec 15]; Available from: https://www.unicef.org/uganda/reports/multidimensional-child-poverty-and-deprivation-uganda-report-volume-one

30. Dowhaniuk N. Exploring country-wide equitable government health care facility access in Uganda. Int J Equity Health [Internet]. 2021 Dec 1 [cited 2022 Apr 6];20(1):1–19. Available from: https://equityhealthj.biomedcentral.com/articles/10.1186/s12939-020-01371-5

